# A high-density 3-dimensional culture model of human glioblastoma for rapid screening of therapeutic resistance

**DOI:** 10.1101/2022.10.22.22281352

**Authors:** J Brown, M Zaben, C Ormonde, F Sharouf, R Spencer, H Bhatt, FA Siebzehnrubl, WP Gray

## Abstract

Glioblastoma is among the most lethal cancers, with no known cure. A multitude of therapeutics are being developed or in clinical trials, but currently there are no ways to predict which patient may benefit the most from which drug. Assays that allow prediction of the tumor’s response to anti-cancer drugs may improve clinical decision-making. Here, we present a high-density 3D primary cell culture model for short-term testing from resected glioblastoma tissue that is set up on the day of surgery, established within 7 days and viable for at least 3 weeks. High-density 3D cultures contain tumor and host cells, including microglia, and retain key histopathological characteristics of their parent tumors, including proliferative activity, expression of the marker GFAP, and presence of giant cells. This provides a proof-of-concept that 3D primary cultures may be useful to model tumor heterogeneity. Importantly, we show that high-density 3D cultures can be used to test chemotherapy response within a 2– 3-week timeframe and are predictive of patient response to Temozolomide therapy. Thus, primary high-density 3D cultures could be a useful tool for brain cancer research and prediction of therapeutic resistance.

## Introduction

Glioblastoma (GBM) is the most lethal type of brain cancer in adults, with a median survival of less than 2 years using the most advanced therapies (Stupp et al., 2015). However, there are no curative therapies available, and consequently the 5-year survival rate of these cancers remains below 5% (Krex et al., 2007). Standard of care for most patients consists of maximum safe resection, followed by radio- and chemotherapy (Alexander and Cloughesy, 2017). Many tumors show *ab initio* or acquired resistance against the most widely used chemotherapeutic agent, Temozolomide (Temodar®, TMZ) (Weller et al., 2010). TMZ resistance has been linked to MGMT promoter methylation status (Combs et al., 2011), but recent evidence indicates that MGMT methylation status may not be predictive of TMZ response in all gliomas. For example, the CATNON trial found no benefit of combined TMZ and radiotherapy versus radiotherapy alone in glioblastoma, *IDH* wild type patients, and while MGMT methylation status was predictive of patient survival it did not predict response to TMZ (Tesileanu et al., 2022). The pathobiology of tumor resistance to TMZ and other chemotherapeutics has been linked to the presence of GBM cancer stem cells (CSCs) (Bao et al., 2006; Siebzehnrubl et al., 2013) and/or the acquisition of stem cell traits in GBM cells (Dirkse et al., 2019).

Molecular diagnosis is now included as part of the standard diagnostic repertoire for GBM (Louis et al., 2021), but to exploit molecular targets for individualized therapies, predictive biomarkers for sensitivity or resistance to targeted therapies are needed. Furthermore, without broad-scale, comprehensive predictive testing of therapeutic resistance or sensitivities of patient tumors, options for clinical decision-making remain limited. There is an urgent need for scalable testing that predicts therapeutic sensitivity of patient tumors in time for initiation of chemotherapy to implement more precision medicine-based approaches in clinical management of GBM.

Here, we describe a high-density 3D *in vitro* primary tumor tissue culture system that is established within 7 days following tumor resection surgery and that recapitulates pathological hallmarks from both tumor and the microenvironment. These primary, high-density 3D cultures can be used to test resistance or sensitivity to chemotherapeutic agents providing a result within 2-3 weeks post-surgery, as we demonstrate with the well-known chemotherapeutic Temozolomide. We show that high-density cultures predict response to Temozolomide therapy, indicating their potential relevance to inform personalized clinical decision-making.

Our data support that the high-density culture method described here is of potential benefit for research and clinical testing, providing an accurate reflection of GBM and host cells that can provide readouts in a clinically relevant timeframe.

## Material and Methods

### Sample collection and tissue culture

GBM tumor samples were collected from patients undergoing surgical resection who gave informed consent for tissue donation through the Welsh Neuroscience Research Tissue Bank approvals process (WNRTB; Ref 19/WA/0058) and all experiments conformed to the Human Tissue Act. Suitable patients were identified by clinical staff at the University Hospital of Wales. Patient data, histological images and reports were anonymized by NHS staff. Clinical follow up data were collected by authors HB and RJS in August 2021 for the patients from whom high density 3D GBM cultures were generated. Data were collected with respect to: (i) MGMT promoter methylation status reported by neuropathology, (ii) oncological therapy received following surgical resection (concurrent temozolomide and radiotherapy followed by adjuvant temozolomide), (iii) progression-free survival (PFS) and (iv) overall survival (OS). Clinical data were analyzed using SPSS statistics 27 (IBM).

Upon collection of tissue samples were immediately placed in 30 ml Gey’s balanced salt solution (Sigma Aldrich) with 10μM (+) MK-801 maleate (Abcam), on ice. Tissue samples were mechanically (McIlwain tissue chopper) and enzymatically digested (DMEM, with 1 μl/ml MK-801 and 2mg/ml papain (Sigma)) at 37°C and 5% CO2 for 45 mins. The cell solution was then strained through 100 and 40 um cell strainers (VWR) to isolate single cells. Cells were centrifuged at 240g for 5 minutes and resuspended at 2.5×10^8^ cells/ml in serum-free media (96% neurobasal A (Life Technologies), 2% B27 (Life Technologies), 1% glutaMAX (Life Technologies) and 1% penicillin/streptomycin (Life Technologies)). 1200 μl media was added to each well of a 6 well plate. Semi-permeable well inserts (Millipore) were placed within each well, and three PTFE confetti disks (HepiaBiosciences) added to each insert. 5 μl of cell suspension was pipetted onto each confetti disk. High-density cultures were incubated at 37ºC and 5% CO2 throughout. 75% media was changed every 2-3 days.

### Immunofluorescence staining

High-density cultures were removed with their confetti to a 48 well plate for fixation (4% paraformaldehyde, 30 minutes) and subsequent immunostaining. Cultures were fixed after 14 days in vitro (DIV) unless otherwise specified. For EdU detection, cultures were exposed to 10 μM EdU for 4 hours before fixation. EdU was then detected using a 647 nm azide using the kit protocol (Click-it EdU imaging kit, Invitrogen). For immunofluorescence staining, cultures were blocked for 30 minutes in PBS-T with 3% donkey serum, then incubated with primary antibodies in the same solution overnight. The following primary antibodies were used: mouse anti b-III tubulin (TuJ1, 1:500, biolegend), rabbit anti Iba1 (1:2000 Wako), rat anti GFAP (1:500, Invitrogen), mouse anti IDH-1 R132H (1:100, Dianova), rabbit anti Nestin (1:500, abcam), mouse anti CD68 (1:500, biolegend), rabbit anti Sox2 (1:500, abcam). Cultures were then washed with PBS-T before incubation with appropriate secondary antibodies in 3% donkey serum in PBS-T (1:500) for 1 hour. Cultures were washed once in PBS-T then incubated with 0.5 ug/ml DAPI for 5 minutes, followed by three further 5 min washes in PBS-T. The confetti discs were then carefully removed from their wells using tweezers, and applied cell-side up to a microscope slide. A droplet of Mowiol mounting solution was placed on top of each culture and a cover slip was applied to the slide.

### Temozolomide treatment

Temozolomide (TOCRIS) was applied to high-density cultures at a range of doses (5, 50, 100, 500 μM) for most samples, and a fixed dose of 100 μM to cultures obtained from regional sampling. Temozolomide was added to the media at 9 DIV. To assess cell proliferation after Temozolomide treatment, cultures were treated with EdU at 14 DIV for 4 hours before fixation. EdU was detected as described above. Concentration-effect curves were generated based on the fraction of EdU-positive cells compared to DMSO treated controls using a 4-parameter logistic equation as described (Eyupoglu et al., 2005).

### Image acquisition and data analysis

Images were taken using an upright fluorescence microscope (Leica DM6000B) or a confocal microscope (Zeiss LSM710). Z-stack confocal images were combined into a maximum-intensity projection. Images were analyzed in ImageJ. DAPI cell counts were performed using an automated process in ImageJ, and other cell counts were performed manually. Cell processes were measured using the distance measurement tool in ImageJ, and microglial clusters were measured using the multiple regions of interest tool.

### Statistical analysis

Statistical testing was carried out using GraphPad Prism 8. Unless otherwise stated, t-tests were used for comparison of two groups, and one-way ANOVA was used for comparison of three or more groups. Unless otherwise specified, data are presented as mean ± SEM.

## Results

### Patients’ Clinical Data

High density 3D cultures were generated from tumor samples from 47 patients with a pathological diagnosis of GBM. We successfully established high density 3D cultures from all GBM patients, with cultures GBM1-GBM28 used for optimizing tissue processing, media formulation and plating density, while cultures GBM29-GBM49 were used for experiments. 3 patients/cultures were excluded due to a final diagnosis other than GBM (two had metastatic disease and one had a grade III oligodendroglioma). Full survival and progression data were available for all patients, however oncological treatment data were missing for one patient. **Table 1** summarizes individual patients’ data. Median age at diagnosis was 65 years (range 38 – 77) and 60% were male.

**Table 1:** Clinical information for all patients and specimens involved in this study.

| Sample number | Sex | MGMT methylation status | IDH1 mutation status | Concurrent TMZ and radiotherapy | Cycles of adjuvant TMZ | Treatment after progression | Progression-free survival | Overall Survival |
| --- | --- | --- | --- | --- | --- | --- | --- | --- |
| GBM1 | M | M | WT | Y | 4 | Nil | 17.1 | 17.1 |
| GBM2 | F | M | WT | Y | 0 | Nil | 12.9 | 12.9 |
| GBM3 | M | Uncertain | WT | Y | 0 | Further surgery | 14.8 | 21.1 |
| GBM4 | M | M | WT | Y | 6 | TMZ | 25.5 | 30.2 |
| GBM5 | F | ND | WT | Y | 6 | PCV and carboplatin | 11.8 | 24.7 |
| GBM6 | M | M | WT | Y | 0 | Nil | 6.0 | 6.0 |
| GBM7 | M | U | WT | Y | 5 | PCV | 10.1 | 12.5 |
| GBM8 | F | M | WT | Y | 6 | TMZ | 25.2 | 46.0 |
| GBM9 | M | M | WT | Y | 6 | “Second-line” chemotherapy | 13.5 | 19.8 |
| GBM10 | M | U | WT | Y | 6 | Further surgery | 20.9 | 27.0 |
| GBM11 | M | M | WT | Missing | Missing | Missing | 7.8 | 7.8 |
| GBM12 | F | ND | WT | N | 0 | Nil | 9.3 | 9.3 |
| GBM13 | M | ND | WT | Y | 2 | Further surgery | 16.8 | 39.3 |
| GBM14 | F | U | WT | Y | 6 | Nil | 17.3 | 25.5 |
| GBM15 | M | U | WT | N | 0 | Nil | 1.2 | 1.2 |
| GBM16 | F | M | WT | Y | 6 | Nil | 42.8 | 42.8 |
| GBM18 | M | M | WT | Y | 6 | Nil | 17.3 | 20.8 |
| GBM19 | F | U | WT | Y | 3 | Nil | 6.0 | 19.4 |
| GBM20 | F | M | WT | Y | 6 | Nil | 12.6 | 14.4 |
| GBM21 | M | U | WT | Y | 3 | Nil | 13.2 | 13.2 |
| GBM22 | F | M | WT | Y | 6 | TMZ, PCV | 28.6 | 39.2 |
| GBM2<br>3 | M | M | WT | Y | 6 | Further surgery | 17.0 | 20.5 |
| GBM2<br>4 | M | M | WT | Y | 0 | Nil | 7.3 | 12.1 |
| GBM2<br>5 | M | M | WT | Y | 6 | Further surgery | 21.8 | 25.0 |
| GBM2<br>6 | M | U | WT | Y | 3 | Nil | 5.7 | 5.7 |
| GBM2<br>7 | F | U | WT | Y | 6 | Lomustine | 9.3 | 15.7 |
| GBM2<br>8 | M | M | WT | Y | 6 | Further surgery | 8.0 | 19.8 |
| GBM2<br>9 | M | M | WT | Y | 6 | Further surgery | 20.3 | 24.1 |
| GBM3<br>0 | F | M | WT | N | 0 | TMZ (initial PCV) | 23.3 | 32.4 |
| GBM3<br>1 | F | U | WT | Y | 0 | Lomustine | 22.6 | 35.2 |
| GBM3<br>2 | M | M | WT | Y | 6 | PCV | 15.1 | 20.7 |
| GBM3<br>3 | F | M | WT | Y | 6 | PCV | 10.5 | 13.3 |
| GBM3<br>4 | F | U | WT | Y | 0 | Nil | 6.3 | 12.8 |
| GBM3<br>5 | M | M | WT | Y | 4 | Nil – but on clinical trial throughout | 22.1 | 28.8 |
| GBM3<br>6 | F | U | WT | Y | 1 | TMZ | 6.5 | 18.2 |
| GBM3<br>7 | M | U | WT | Y | 5 | Lomustine | 10.0 | 11.4 |
| GBM3<br>9 | M | U | WT | Y | 0 | Nil | 4.3 | 5.1 |
| GBM4<br>0 | M | U | WT | Y | 6 | Lomustine, carboplatin | 13.2 | 20.0 |
| GBM4<br>1 | F | M | MUT | Y | 6 | Nil | 27.6 | 27.6 |
| GBM4<br>2 | M | U | WT | Y | 1 | Nil | 6.4 | 6.4 |
| GBM4<br>3 | M | U | MUT | Y | 4 | Nil | 22.4 | 27.5 |
| GBM4<br>4 | F | U | WT | Y | 4 | PCV | 13.1 | 19.0 |
| GBM4<br>5 | F | M | WT | Y | 6 | Nil | 27.3 | N/A |
| GBM4<br>6 | M | U | WT | Y | 6 | PCV | 11.1 | 21.0 |
| GBM4<br>7 | M | U | WT | Y | 6 | PCV | 9.5 | 14.2 |
| GBM4<br>8 | M | M | WT | Y | 6 | TMZ | 22.5 | N/A |
| GBM4<br>9 | F | M | WT | Y | 5 | Nil | 13.2 | 19.7 |

Summary of clinical information
| Variable |  | Number / Median | %age of total / total range |
| --- | --- | --- | --- |
| Age (years) |  | 65 | 38 – 77 (IQR 56 – 69) |
| Male sex |  | 28 | 59.6 % |
| MGMT promoter methylation status | Methylated | 24 | 51.1 % |
|  | Unmethylated | 19 | 40.4 % |
|  | Unknown | 4 | 8.5 % |
| IDH1 mutation status | R132H mutation | 2 | 4.3 % |
|  | Wild-type | 45 | 95.7 % |
| Concurrent TMZ and radiotherapy received |  | 43 | 91.5 % |
| Adjuvant TMZ (number of cycles) | 0 | 10 | 21.3 % |
|  | 1 | 2 | 4.3 % |
|  | 2 | 2 | 4.3 % |
|  | 3 | 3 | 6.4 % |
|  | 4 | 3 | 6.4 % |
|  | 5 | 3 | 6.4 % |
|  | 6 | 22 | 46.8 % |
|  | >6 | 1 | 2.1 % |
| Progression-free survival (months) |  | 13.2 | 1.2 – 42.8 (IQR 9.3 – 21.8) |
| Overall survival (months) |  | 19.8 | 1.2 – 46.0 (IQR 12.9 – 27.5) |

24 patients’ tumors (55.8%) had methylated MGMT promoters. 19 patients had unmethylated tumors and MGMT promoter methylation status was missing for four patients. There was no significant difference in mean age at diagnosis between methylated and unmethylated patients (t-test, p=0.453). Two tumors were classified as grade 4 astrocytoma, IDH-mutant (R132H) (Louis *et al*., 2021) and the remaining 45 were grade 4 GBM, IDH-wild type. Median PFS was 13.2 months while median OS was 19.8 months (**Fig. 1A,B**). Two patients were still alive at the time of manuscript writing.

**Figure 1:**
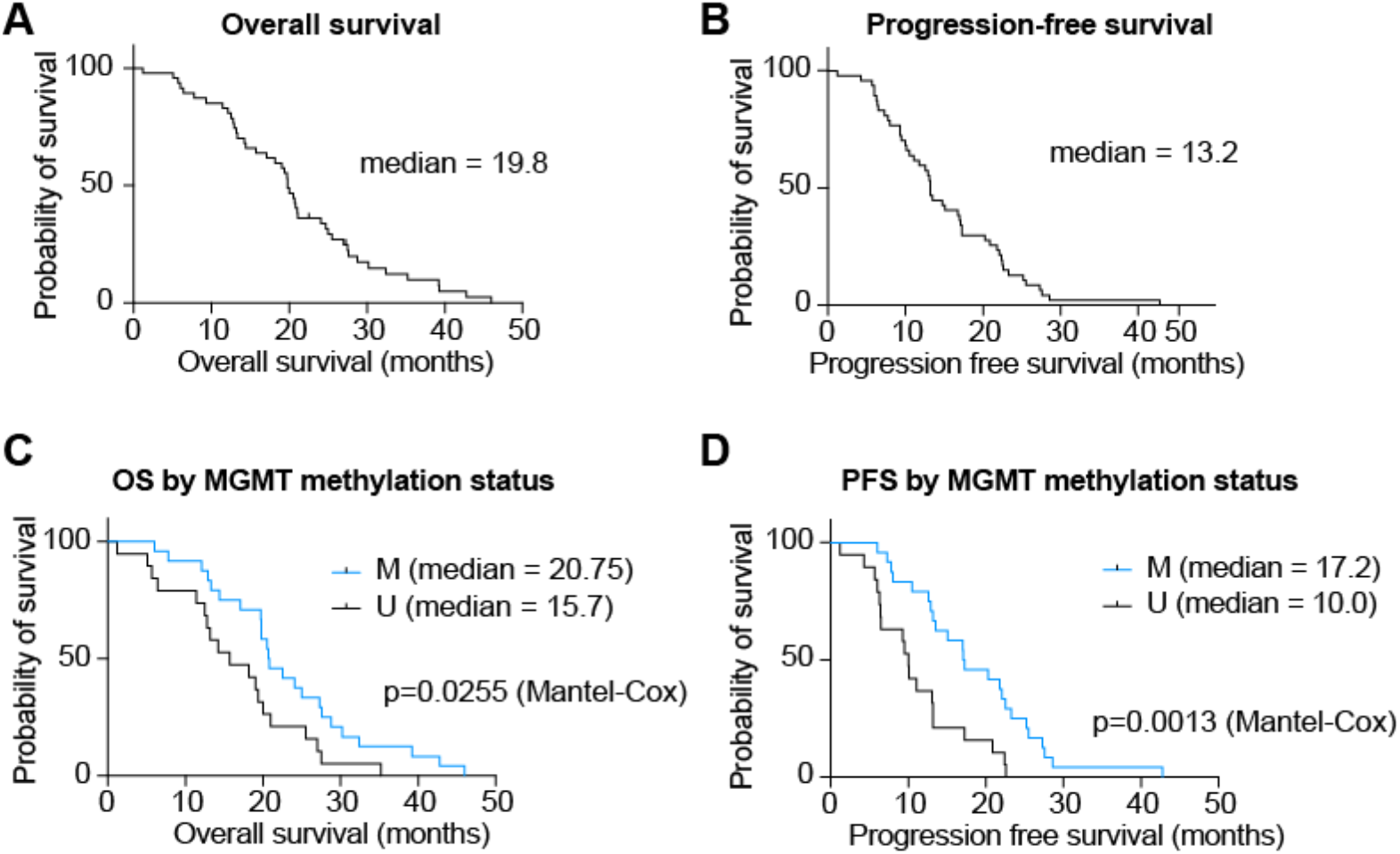
Kaplan-Meier survival curves for patients included in this study. **(A)** Overall survival. **(B)** Progression-free survival. **(C)** Overall survival according to MGMT methylation status. **(D)** Progression-free survival according to MGMT methylation status.

91.5% of patients received the standard of care six weeks of concurrent temozolomide (TMZ) and radiotherapy after their surgical debulking. Patients received a median of 5.5 cycles of further adjuvant TMZ. Six patients did not receive any TMZ (see **Table 1** for details). Patients with methylated tumors received significantly more cycles of adjuvant TMZ than their unmethylated counterparts (median 6 vs 3 cycles, Mann-Whitney *U* test, p = 0.027).

In Kaplan-Meier survival analysis (**Fig. 1C,D**), the methylated group had a significant survival benefit compared to unmethylated patients in both PFS (median 17.2 vs 10 months, Mantel-Cox log rank χ ^2^ = 10.33, p = 0.0013) and OS (median 20.75 vs 15.7 months, Mantel-Cox log rank χ ^2^ = 4.989, p = 0.0255).

### Characterization of high-density 3D cultures from human GBM

To establish an *in vitro* system that contains GBM cells as well as local microenvironmental cells and that is suitable for screening tumor cell response to therapeutics in a short timeframe, we adapted a method that we previously established in primary human hippocampal cultures (Zaben et al., 2021). To generate high-density cultures, for each culture 5 μl of a single cell suspension of primary GBM cells at a density of 2.5 × 10^8^ cells/ml were plated on a PTFE membrane. High-density cultures were grown at the media-air interface using a defined serum-free medium without additional mitogens (e.g., EGF, FGF2; **Fig. 2A,B**). High-density cultures show an initial consolidation phase over the first week in culture that is characterized by a significant decreased in cell numbers, followed by a stable culture period thereafter with no significant changes in cell numbers between time points (**Fig. 2C**). Likewise, the labelling index of cell proliferation in the high-density cultures (i.e., cell cycle rate), defined by the ratio of EdU+Ki67+ over total Ki67+ fraction, remains stable over the culture period. A lower labelling index at 21 days in culture could indicate eventual decrease of proliferation rates, but this was not found significant (**Fig. 2D**). Thus, high-density cultures are established at the day of surgery and have consolidated into a usable *in vitro* system within 7 days after patient resection.

**Figure 2:**
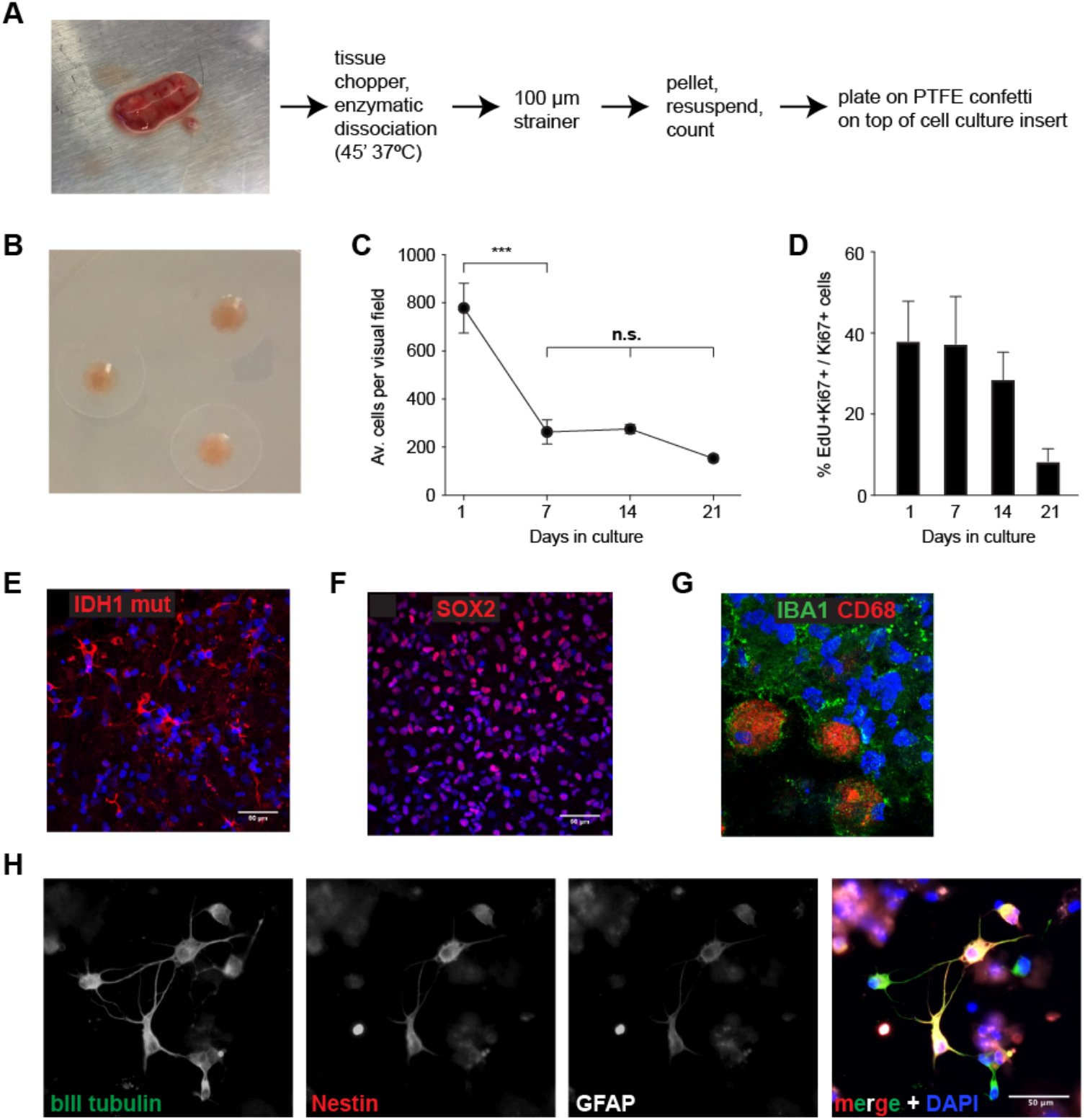
Characterization of high-density cultures. **(A)** Schematic of human high-density culture preparation. Tissue from surgical resections (left image) is mechanically and enzymatically dissociated, passed through a 100 μm strainer to remove debris, and centrifuged before plating a defined number of cells (see Material and Methods) onto a PTFE membrane placed on top of a cell culture insert at the liquid-air interface. **(B)** Image shows 3 established high-density cultures on top of PTFE membranes. **(C)** After an initial consolidation phase (day 1-7), the number of cells within high-density cultures remain stable over at least 3 weeks (n=3 cultures; *** p<0.001, n.s. not significant). **(D)** Labelling index shows active and stable proliferation within high-density cultures over 2 weeks that eventually slows around 3 weeks after plating. **(E)** Immunofluorescence staining for IDH1-R132H demonstrates that high-density cultures contain both tumor and non-tumor cells. Scale bar, 50 μm. **(F)** Immunofluorescence staining for SOX2 highlights presence of cancer stem cells in high-density cultures. Scale bar, 50 μm. **(G)** Immunofluorescence staining confirmed presence of microglia (IBA1) of different activation states (CD68) in high-density cultures. **(H)** GBM cells in high-density cultures frequently co-express neuronal (bIII tubulin), progenitor (Nestin) and astrocytic (GFAP) markers. Scale bar, 50 μm (applies to all images).

We next characterized the cell types present in high-density cultures. To conclusively label tumor cells within high density cultures, we exploited the IDH1 mutation which is typically shared in >95% of tumor cells as it occurs early during tumorigenesis (Capper et al., 2009). We selected a patient with IDH1-mutant (R132H) astrocytoma to assess the prevalence of tumor cells in high-density cultures (**Fig. 2E**). IDH1-R132H staining confirmed presence of tumor cells, but also showed a significant fraction of non-tumor cells in high-density cultures. We further evaluated tumor and non-tumor cell populations in high density cultures in IDH wild type GBM patients. To assess presence of GBM cancer stem cells we used immunofluorescence staining for the cancer stem cell marker SOX2 (**Fig. 2F**). To evaluate non-tumor cell populations in high-density cultures we focused on microglia, which constitute the most frequent cell type within GBM (Glass and Synowitz, 2014). Co-staining for IBA1 and CD68 demonstrated presence of microglia in high-density cultures and revealed heterogeneity of microglial activation stages (**Fig. 2G**). To further characterize GBM cells within high-density cultures, we co-stained for the early neuronal marker tubulin beta III (TuJ1), the stem/progenitor marker Nestin and the astrocyte marker GFAP (**Fig. 2H**). Most GBM cells expressed all three markers, although there was some heterogeneity among the intensity of individual marker expression.

Our findings show that high-density 3D cultures can be established within 7 days of surgical resection, are viable over at least 3 weeks, and contain tumor and non-tumor cells.

### High-density cultures share histopathological features of parent tumors

Next, we determined to which degree high-density cultures reflect the pathology of the original patient tumor (**Table 2**). For this, we compared immunohistochemistry and H&E staining of patient material taken for histopathological diagnosis with immunofluorescence staining of high-density cultures. We found that high-density cultures expressed histopathological marker GFAP in similar levels as their parent tumors (**Fig. 3A**). High-density cultures also showed EdU uptake at comparable levels to proliferation levels in their parent tumors (**Fig. 3B**). The presence of giant cells is a hallmark of a subgroup of GBMs (Louis *et al*., 2021), a finding which was mirrored in high-density cultures. Cultures from 3 patients where giant cells were confirmed by histopathology showed giant nuclei as well. High-density cultures from 12 patients where no giant cells were detected in material sent for neuropathological diagnosis did not contain giant cells (**Fig. 3C**).

**Table 2:** Comparison of high-density 3D cultures and patient pathological features. A summary of relevant data from pathology reports of patient GBM samples, showing a range of Ki-67% levels. Values are shown as reported by a pathologist. These have been allocated into three categories: high, medium, low. High-density culture EdU positivity levels are also reported as high, medium, or low.

| Sample | Ki-67 pathology | Pathology results | Culture results |
| --- | --- | --- | --- |
| GBM30 | Up to 50% | HIGH | HIGH |
| GBM31 | <1% | LOW | MED |
| GBM33 | 60% | HIGH | HIGH |
| GBM34 | 20% | MED | LOW |
| GBM37 | 20-25% | MED | MED |
| GBM39 | 40% | HIGH | HIGH |
| GBM43 | Low, but areas of 20-40% | LOW | LOW |
| GBM44 | 30% | MED | MED |
| GBM45 | 30% | MED | LOW |
| GBM46 | Up to 25% | MED | LOW |
| GBM47 | Up to 30% | MED | HIGH |
| GBM48 | 40% | HIGH | LOW |
| GBM49 | 25% | MED | LOW |

**Figure 3:**
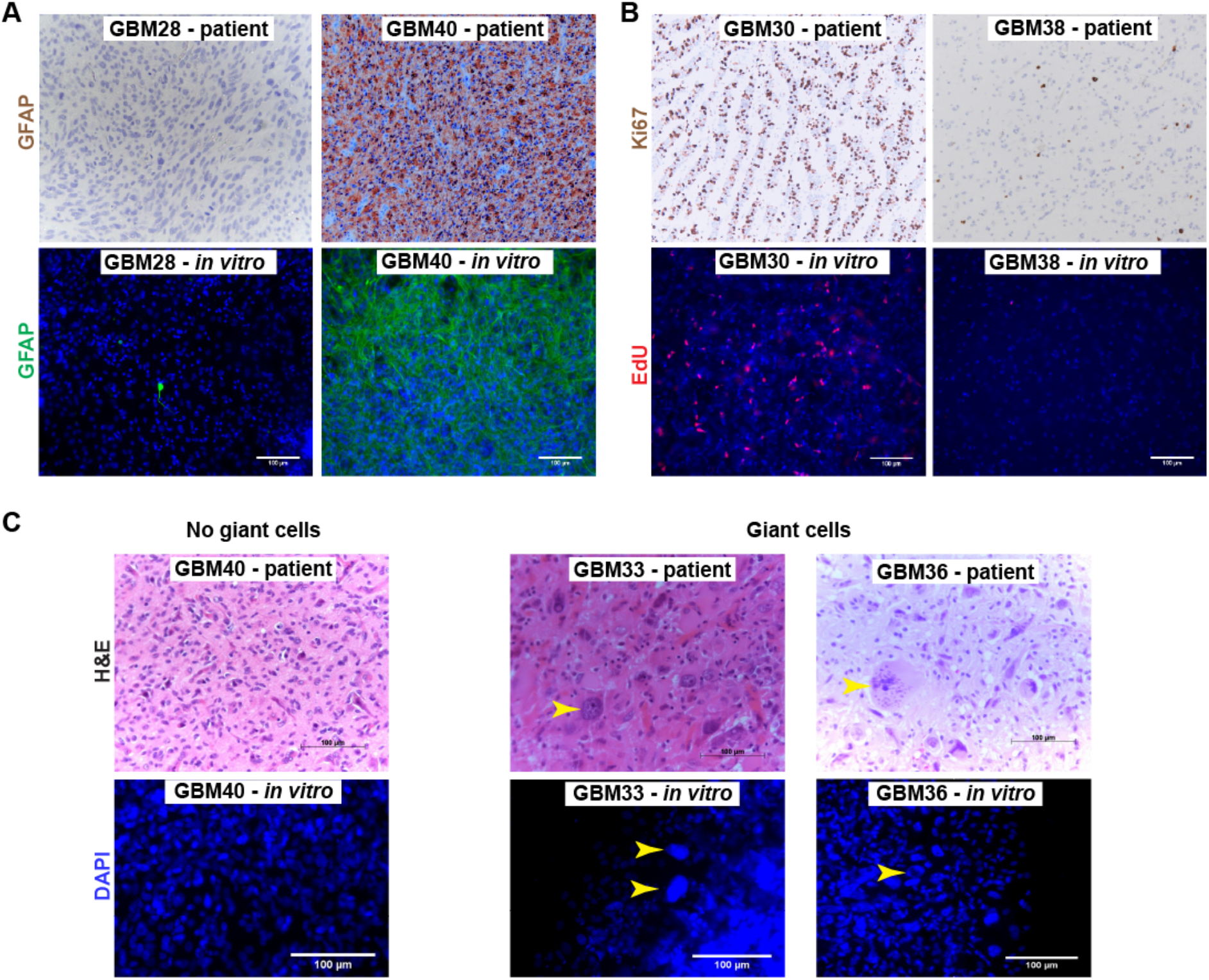
High-density share neuropathological features of parent tumors. Established high-density cultures retain histopathological hallmarks of parent tumors, including immunoreactivity for GFAP **(A)**, frequency of proliferating cells **(B)** and the presence of giant cells **(C)**. Depicted are representative images of parental tumors (top row) and the corresponding high-density cultures (bottom row). Arrowheads indicate giant cells. Scale bars, 100 μm.

Together, this shows that high-density cultures recapitulate histopathological characteristics of their parent tumors. We next asked whether high-density cultures also reflect treatment response of patient tumors.

### High-density 3D cultures exhibit chemoresistance profiles of parent tumors and predict patient response to chemotherapy

Because high-density cultures shared histopathological features with the patient material, we wanted to determine whether these cultures could also mirror treatment response characteristics of their parent tumors, and whether these cultures could be used to predict response to chemotherapy. Therefore, we assessed cellular response of high-density cultures after exposure to increasing concentrations of the standard chemotherapeutic TMZ compared to DMSO-treated controls by quantifying short-term EdU incorporation, which is proportional to proliferation rates. Experimenters conducting dose-response testing of TMZ were blinded to the neuropathological and MGMT promoter methylation-status of the parent tumors. We noted that some high-density cultures were sensitive to TMZ, with an IC_50_ of EdU incorporation in the μM range (**Fig. 4A**), whereas other cultures showed TMZ resistance with mM IC_50_ (**Fig. 4B**). We were able to establish whether a culture was TMZ sensitive or resistant within 14 days of surgical tumor resection.

**Figure 4:**
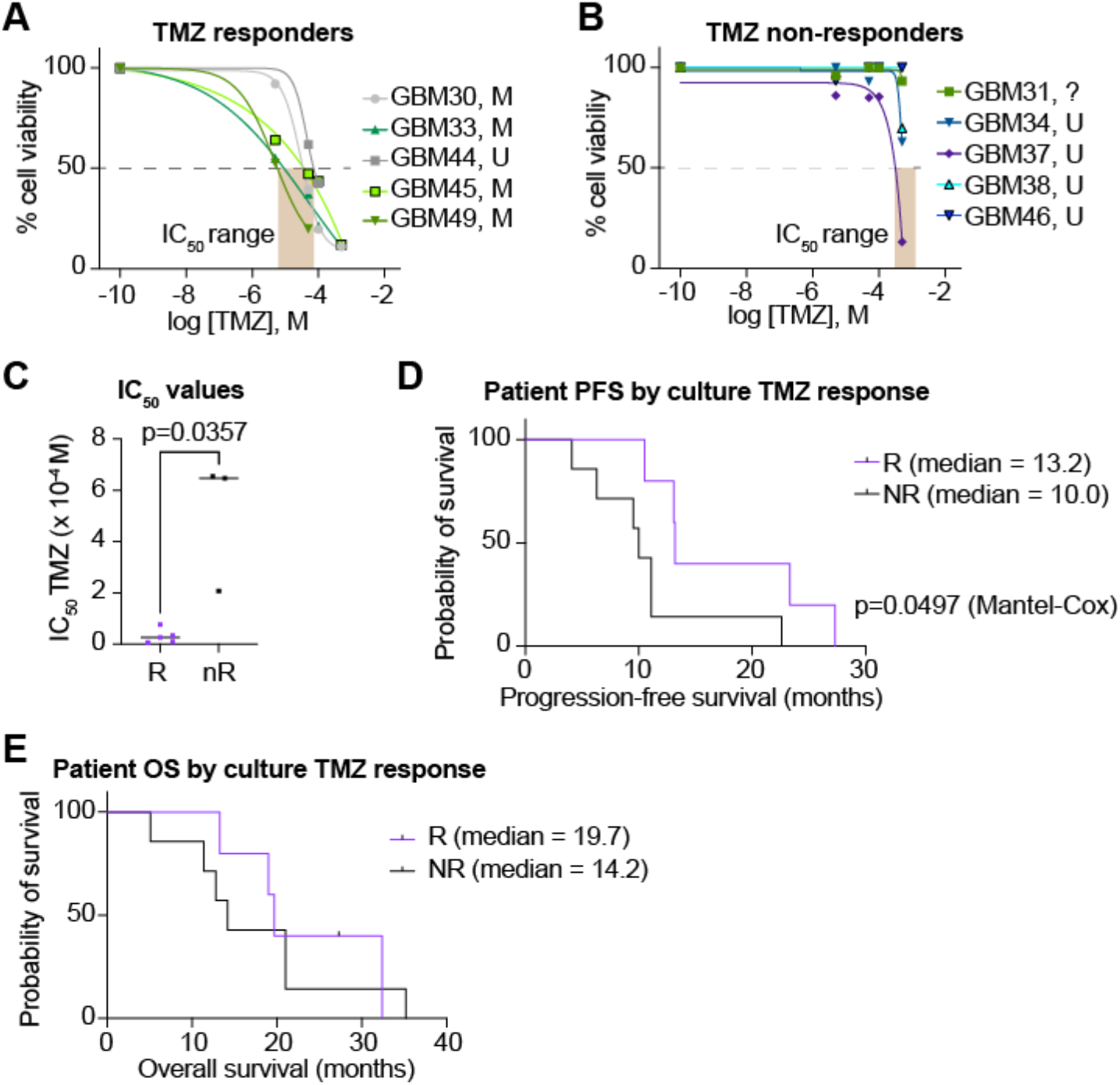
Chemoresistance profiles of high-density cultures. Cell proliferation rates based on short-term EdU incorporation of high-density cultures after Temozolomide (TMZ) treatment reflects patient MGMT methylation status (**A**, TMZ sensitive cultures; **B**, TMZ resistant cultures; n=5 replicates per culture; greyed area indicates IC_50_ concentration ranges). TMZ resistance correlates with MGMT promoter methylation status of parental tumor (M: methylated, U: unmethylated, ?: unknown). **(C)** Comparison of IC_50_ values from TMZ-sensitive (responding, R) and TMZ-resistant (non-responding, nR) cultures. **(D)** Progression-free survival of patients stratified according to high-density culture response (responding: R.; non-responding: NR) **(E)** Overall survival of patients stratified according to high-density culture response (responding: R.; non-responding: NR).

When comparing TMZ responses of high-density cultures post-hoc with MGMT promoter methylation status, we found that TMZ sensitive cultures were almost exclusively derived from patients with MGMT promoter methylation, whereas TMZ resistant cultures were mostly derived from patients with unmethylated MGMT promoters (**Fig. 4A,B**). MGMT promoter methylation is frequently associated with a hypermethylated phenotype (G-CIMP) in GBM (Brennan et al., 2013) and with prolonged survival in patients treated with TMZ (Aldape et al., 2015). Statistical analysis of IC_50_ values from TMZ sensitive (responding, R) versus TMZ resistant (non-responding, NR) cultures revealed a significant difference (**Fig. 4C**; p=0.0357, Mann-Whitney test).

We next compared the response of high-density cultures with patient outcomes. When patients were stratified according to TMZ response of their corresponding high-density cultures, we found a significant difference in progression-free survival of patients with TMZ-sensitive cultures (responding, R) compared to patients with TMZ-resistant cultures (non-responding, NR) (**Fig. 4D**). This may indicate that patient tumors with TMZ-sensitive cultures showed better response to this chemotherapeutic agent than tumors from patients with TMZ resistant cultures. Of note, we did not find a significant difference in overall survival of patients stratified according to TMZ response of their corresponding high-density cultures (**Fig. 4E**), indicating that high-density cultures are more predictive of immediate response of patient tumors rather than long-term outcomes. This demonstrates a predictive effect of high-density culture sensitivity, established within 14 days of resection, on patient progression-free survival.

## Discussion

There is an urgent need to improve clinical management of patients with GBM. Incredible progress has been made over the last 2-3 decades in understanding molecular determinants of GBM tumor development and progression (Brennan *et al*., 2013; Ceccarelli et al., 2016; Verhaak et al., 2010), as well as the cellular and molecular basis of the heterogeneity of this disease (Neftel et al., 2019; Patel et al., 2014). But so far, little of this progress has been successfully translated into new therapies. The profound heterogeneity of GBM has highlighted that it is unlikely that there will be a ‘one-size-fits-all’ therapy, and some degree of individualized treatment that is somehow tailored to the patient’s tumor is likely to be necessary.

The method described here allows generating hundreds of high-density cultures from an individual patient simultaneously and within 1-2 weeks following surgical resection of tissue. The number of parallel cultures as well as the short timeframe for establishing these cultures positions this system exquisitely for rapid clinical testing of drug sensitivities as shown here. We demonstrate proof-of-concept that a biological readout of TMZ resistance can be obtained within 2-3 weeks after surgery and is predictive of patient response to TMZ chemotherapy as evaluated by PFS. TMZ resistance patterns in high-density cultures from patient tumors complemented standard molecular pathology MGMT promoter methylation testing. While there was no perfect correlation, in most patients MGMT methylation status agreed with TMZ sensitivity. Since MGMT methylation is not predictive in all high-grade gliomas (Tesileanu *et al*., 2022), additional methods that predict tumor response to chemotherapy could help close a gap in clinical decision making.

In addition to similar molecular characteristics, established high-density cultures exhibited cellular and histopathological hallmarks of parent tumors, including GFAP expression, proliferation, and the presence of giant cells. These features render high-density cultures a potentially useful tool to study tumor pathology and biology. Additionally, the presence of key immune cells such as microglia, as well as GBM cancer stem-like cells within high-density cultures indicates their potential for reflecting critical cell-cell interactions in the tumor microenvironment. Therefore, high-density cultures may prove valuable in both research and clinical settings. The rapid set-up and short-term readout from these cultures further enables their broad application across many settings.

The high-density cultures comprise a 3D short-term tissue culture model that includes both GBM and TME immune components. Thus, high-density cultures reflect TME elements that conventional cell culture models and some animal models are missing (e.g., xenograft models lacking immune components) (Robertson et al., 2019). Genetically engineered mouse models of GBM allow orthotopic tumor initiation and progression within a fully intact TME but rely on a defined set of mutations driving tumor growth that only incompletely reflect the diverse and heterogeneous mutational landscape of these cancers (Robertson *et al*., 2019). By contrast, patient-derived xenograft (PDX) models are considered the current gold standard in oncology research and for predicting response to therapy (Koga and Ochiai, 2019). While these models are closer to recapitulating a TME than conventional subcutaneous xenograft models, they are technically demanding, and require immune suppression to prevent graft rejection. Hence, PDX models critically lack immune cell components which can influence cancer progression and response to therapy. PDX models are also not readily amenable to medium-high throughput investigations, given the overall technical challenges that PDX models present. Additionally, the growth rates of GBM mouse models limit their usefulness for predicting individual responses in patients (precision medicine) within a clinically relevant timeframe. These issues further emphasize the need for immune competent GBM models, which fully recapitulate interactions between human tumor cells and the surrounding normal human brain environment. Many contemporary 3D models rely on co-culturing patient derived GBM cells with TME components derived from pluripotent stem cells (e.g., hiPSC), but these will lack a TME that has been educated by the patient tumor. A recent landmark study demonstrated that GBM organoids can be generated and used in a similar timeframe as the high-density cultures described here (Jacob et al., 2020). Thus, our work extends the available experimental toolbox for rapid establishment of GBM cultures that reflect essential hallmarks of the patient tumor, and which are suitable for clinically relevant assays to predict patient response to therapy.

We present a method for primary 3D cultures of GBM tissue that can provide a rapid readout of drug sensitivity. Currently, our findings are limited by the number of patients analyzed, but even in the small cohort available we showed a predictive value of high-density 3D cultures. Furthermore, molecular genetic profiling of primary cell cultures and parent tumors could provide important validation that primary cultures reflect molecular subclasses of parent tumors.

In summary, we describe a rapid and simple method for generating high-density cultures from patient GBM tissue that is amenable to clinical and research settings, allows capturing regional heterogeneity and is predictive of patient therapy response.

## Data Availability

All data produced in the present study are available upon reasonable request to the authors.

## Acknowledgements

The authors would like to thank members of the WPG and FAS labs for critical comments on the manuscript. WPG is funded by NC3Rs project NC/N002423/1. FAS is supported by MRC grant MR/S007709/1. The BRAIN Unit is funded by Welsh Government through Health and Care Research Wales (BRAIN Unit Infrastructure Award, Grant no: UA05 to WPG).

## Declaration

The authors declare no conflict of interest.

## Notes

### Competing Interest Statement

The authors have declared no competing interest.

### Funding Statement

This study was funded by the NC3Rs, the MRC, and Health and Care Research Wales.

### Author Declarations

Research Ethics Committee of Cardiff University gave ethical approval for this work.

